# Mucosal antibody response to SARS-CoV-2 in paediatric and adult patients: a longitudinal study

**DOI:** 10.1101/2021.09.27.21264219

**Authors:** Renee WY Chan, Kate CC Chan, Grace CY Lui, Joseph GS Tsun, Kathy YY Chan, Jasmine SK Yip, Shaojun Liu, Michelle WL Yu, Rita WY Ng, Kelvin KL Chong, Maggie H Wang, Paul KS Chan, Albert M Li, Hugh S Lam

**Affiliations:** Department of Paediatrics, Faculty of Medicine, The Chinese University of Hong Kong, Hong Kong SAR; Laboratory for Paediatric Respiratory Research, Li Ka Shing Institute of Health Sciences, Faculty of Medicine, The Chinese University of Hong Kong, Hong Kong SAR; CUHK-UMCU Joint Research Laboratory of Respiratory Virus & Immunobiology, Department of Paediatrics, Faculty of Medicine, The Chinese University of Hong Kong, Hong Kong SAR; Hong Kong Hub of Paediatric Excellence, The Chinese University of Hong Kong, Hong Kong SAR; Department of Medicine and Therapeutics, Faculty of Medicine, The Chinese University of Hong Kong, Hong Kong SAR; Department of Paediatrics, Prince of Wales Hospital, New Territories, Hong Kong SAR; Department of Microbiology, Faculty of Medicine, The Chinese University of Hong Kong, Hong Kong SAR; Department of Ophthalmology and Visual Sciences, Faculty of Medicine, The Chinese University of Hong Kong, Hong Kong SAR; The Jockey Club School of Public Health and Primary Care, Faculty of Medicine, The Chinese University of Hong Kong, Hong Kong SAR

## Abstract

Conjunctival and nasal mucosal antibody responses in thirty-four paediatric and forty-seven adult COVID-19 patients were measured. The mucosal antibody was IgA dominant. In the nasal epithelial lining fluid (NELF) of asymptomatic paediatric patients, SARS-CoV-2 spike protein 1 (S1) specific immunoglobulin A (IgA) was induced early. Their plasma S1-specific IgG levels were higher than symptomatic patients. More adult with mild disease had NELF S1-specific IgA than those with severe/critical illness. Within the first week of diagnosis, higher S1-specific antibodies in NELF and plasma and lower vial loads were detected in paediatric than adult patients with mild disease. The IgA and IgG levels correlated positively with the surrogate neutralization readout. The detectable NELF ‘neutralizing’ S1-specific IgA in the first week after diagnosis correlated with a rapid decline in viral load. This study highlights the effect of nasal IgA in limiting the SARS-CoV-2 replication and provides complementary information to the serum antibody measurements.

## Main

Severe acute respiratory syndrome coronavirus 2 (SARS-CoV-2) causes Coronavirus disease 19 (COVID-19). ^1^ SARS-CoV-2 interacts with the angiotensin-converting enzyme 2 (ACE2) expressed by the nasal epithelia for entry and infect neighbouring epithelial cells,^2, 3^ while conjunctival goblet cell is suggested to be an alternative portal of entry. ^4, 5^ Therefore, examining the mucosal antibody of COVID-19 patients will allow a more thorough discernment of the viral-host interaction and the underlying immunopathology. Although mucosal immunity plays a major role in SARS-CoV-2 infection, most studies have focused on systemic immunity.^6, 7, 8^ There is a paucity of knowledge on the SARS-CoV-2 specific antibodies on the conjunctival and respiratory mucosa.

Mucosal immunity is achieved by innate and acquired immune responses.^9, 10^ Viral antigens acquired locally in the conjunctival and nasal epithelium are processed in the conjunctiva-associated lymphoid tissue (CALT)^11^ and nasopharyngeal-associated lymphoid tissue (NALT) respectively.^12^ Meanwhile, these lymphoid tissues generate IgA-producing mucosal B cells that express homing receptors for efficient trafficking to the mucosal effector site.^13, 14^ Secretory IgA is a potent dimeric IgA found on mucosal surfaces,^15^ and provides broader protection due to its higher avidity.^16^ It is responsible for agglutinating and neutralizing the virus on the respiratory tract, in the respiratory cell and within the lamina propria beneath the epithelium. The dimeric form is found to be fifteen fold more potent than its monomeric counterpart in plasma.^17^ Moreover, it provides effective immunity against infection when compared with its IgG isotype in a monoclonal antibody study of Mab362, ^18^ and in the investigation of antibody neutralization in a natural disease course of SARS-CoV-2 infection.^19^

The early and intense induction of serological IgA in COVID-19 patients has been reported.^7^ Sterlin *et al* documented that the first wave of circulating IgA-expressing plasmablasts precedes the IgG-expressing cells.^19^ Together with the fact that IgA is potent in virus neutralization, IgA contributed significantly in the early phase of the infection. Therefore, we hypothesized that early detection of SARS-CoV-2 specific IgA in the mucosal fluid would correlate with a lower viral load and milder symptoms. Cervia *et al* reported that mild disease or low antigen exposure might stimulate mucosal SARS-CoV-2–specific IgA response, which could be accompanied by the absence, presence or delayed systemic virus-specific IgA production.^6^ Such pattern appears to be particularly prevalent in younger individuals and might explain why children commonly present with asymptomatic or mild SARS-CoV-2 infection. However, this hypothesis requires further supportive evidence from longitudinal studies in both children and adult patients.

Our study aimed to evaluate the longitudinal SARS-CoV-2 specific antibody levels in the mucosa and their neutralising effect to address the research gap. A major limitation in mucosal immunity research is the inaccuracy of the results due to mucosal sample collection methods, such as, the nasal swabs or irrigation methods, which are subjected to low sample yield and inconsistency of the dilution effect.^20, 21^ In our study, conjunctival fluid (CF) samples were collected with a technique similar to the Schirmer’s test^22, 23^ while the nasal epithelial lining fluid (NELF) samples were collected by nasal strips.^24, 25^ These methods are standardised and exhibit extended sample stability even when stored at room temperature,^26^ which ensure sample validity.

The mucosal antibody kinetics described in this longitudinal study conducted in paediatric and adult COVID-19 patients identified the differential SARS-CoV-2 specific antibody levels detected in asymptomatic and symptomatic patients. It also highlighted the association between early and intense levels of the nasal SARS-CoV-2 specific immunoglobulin A (IgA) with the more rapid decline in viral load in patients. This study may shed light on the role of mucosal immunity in reflecting SARS-CoV-2 exposure and protection against transmission to guide future diagnostics and vaccines development.

## Results

### Demographics of the subjects

Thirty-four paediatric patients and forty-seven adult patients participated in this study. All subjects tested negative to other respiratory pathogens in the multiplex panel during admission. The median age was 12.5 years old (range 6-17) for the paediatric group and 61 years old (range 18-88) for the adult group, with 32% and 39% male subjects in the respective groups (**Figure 1b**). These subjects were assigned with a severity score based on the criteria listed in the WHO living guidance.^27^ Except for one patient who had moderately severe disease, all other paediatric subjects had mild disease, of which 14 were. As there were only two critically ill patients among adult participants, we pooled the severe and critically ill groups together for the final analysis. A total of 145 CF, 454 NELF and 158 plasma samples were collected (**Figure 1b**).

**Figure 1.**
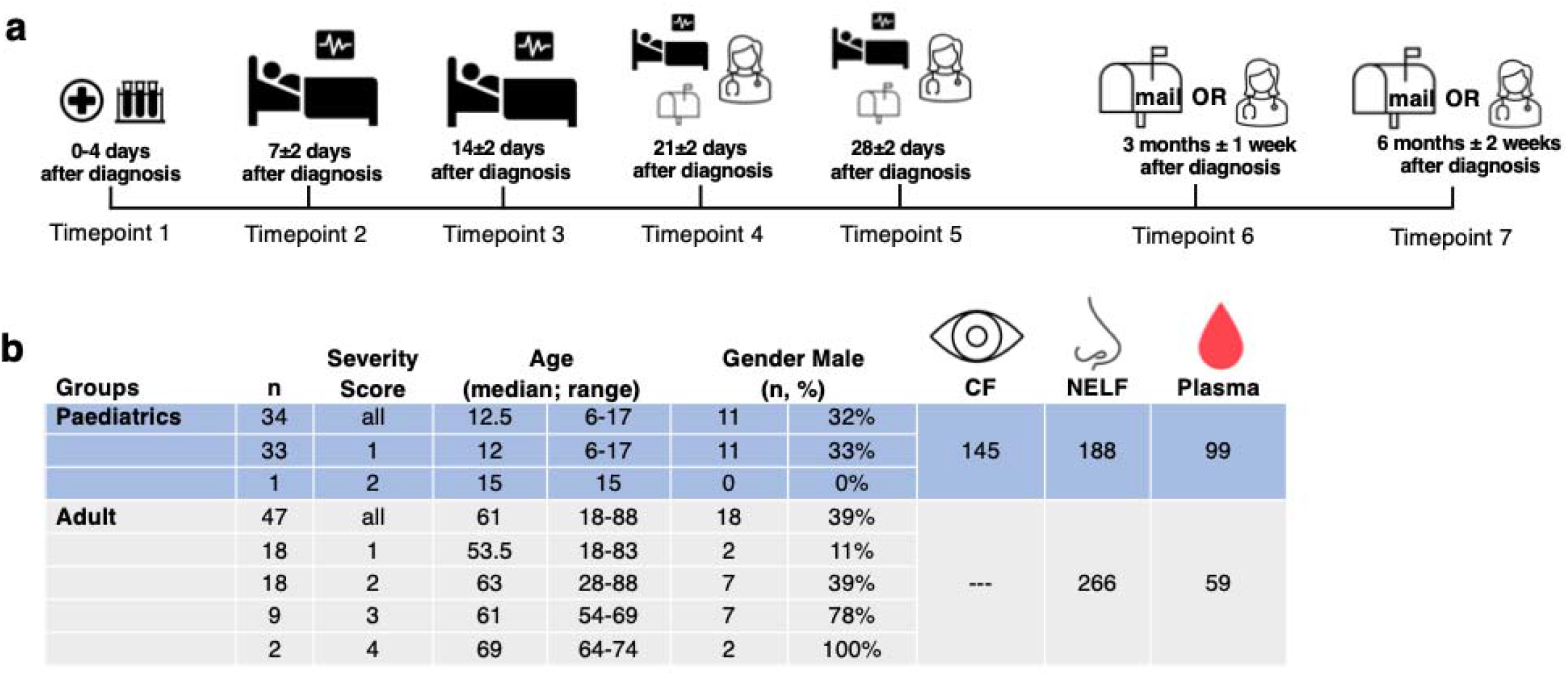
Study design and demographics. **(a)** A longitudinal sample collection from the day of diagnosis (disease onset or the first day of SARS-CoV-2 PCR positive, whichever earlier) to six months post diagnosis was conducted by healthcare workers during hospitalization and follow-up consultation for paediatric patients. Adult patients performed self-collection of NELF samples after being discharged and mailed the samples to the laboratory. **(b)** the number of paediatric and adult subjects, severity score, age, gender and the number of CF, NELF and plasma collected are shown.

### SARS-CoV-2 S1-specific IgA dominated conjunctival and nasal epithelial lining fluids

SARS-CoV-2 S1-specific IgA was detected in 50% of CF, 54% of the NELF and 43% of the plasma samples of the paediatric patients within the first four days of disease diagnosis. S1-specific IgG was not detected in CF at any of the time points (**Figure 2a**). A minority of NELF samples tested positive to S1-specific IgG twelve days after disease diagnosis (**Figure 2b**). In plasma, the S1-specific IgA could be detected earlier than IgG and at a higher level in the first 23 days after diagnosis. In contrast, S1-specific IgG became dominant by 3 months after diagnosis (**Figure 2c**). At six months post-diagnosis, 53% of CF, 45% of NELF remained S1-specific IgA positive, while 64% and 79% of plasma remained S1-specific IgA and IgG positive, respectively.

**Figure 2.**
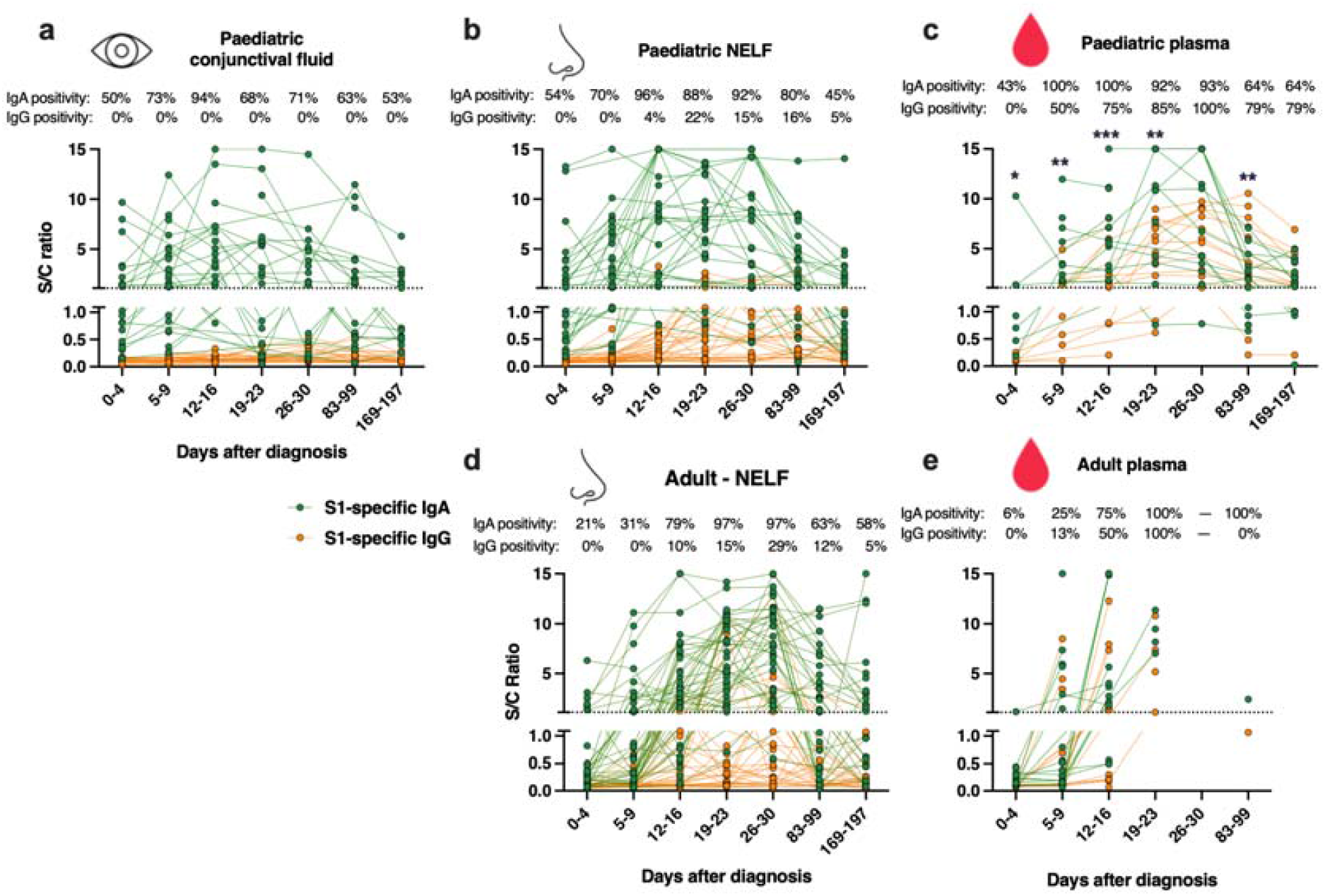
SARS-CoV-2 S1-specific antibody levels in COVID-19 patients. The longitudinal changes of SARS-CoV-2 S1-specific IgA 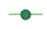 and IgG 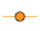 in the **(a)** conjunctival fluid, **(b)** nasal epithelial lining fluid (NELF) and **(c)** plasma of paediatric patients and the **(d)** NELF and **(e)** plasma of adult patients were plotted. Data points above the dotted line (Sample/Calibrator (S/C) ratio ≥ 1.1) are considered as positive, while y=15 indicates the upper detection limit of the assay. The percentages denote the IgA and IgG positivity at each time point. The lines connect data points of the same patients, while the asterisks indicate the statistical differences found between the S/C ratio of S1-specific IgA and IgG in plasma samples by Wilcoxon matched-pairs signed rank test, *: *p* <0.05; **: *p* < 0.01; ***: *p*< 0.005.

In NELF of adult patients, S1-specific IgA was also the dominant isotype (**Figure 2d**). Overall, 21% and 31% of adult NELF collected on 0-4 and 5-9 days post-diagnosis was S1-specific IgA positive. At six-month post-diagnosis, 58% of NELF remained S1-specific IgA positive. However, we did not have plasma samples at this time point to assess the longevity of S1-specific IgA and IgG.

### Symptomatic COVID-19 paediatric patients had a higher level of S1-specific IgA in CF

Twenty-nine paediatric patients provided serial CF samples for longitudinal measurements and 93% (27/29) of the patients had S1-specific IgA in their CF in at least one time point. Symptomatic children had higher levels of S1-specific IgA on 12-16 and 26-30 days post-diagnosis (*p* = 0.0140 and 0.0246, respectively (**Figure 3a, Table 1**) while no S1-specific IgG was detectable in any of the available samples (**Figure 3b**).

**Table 1.**
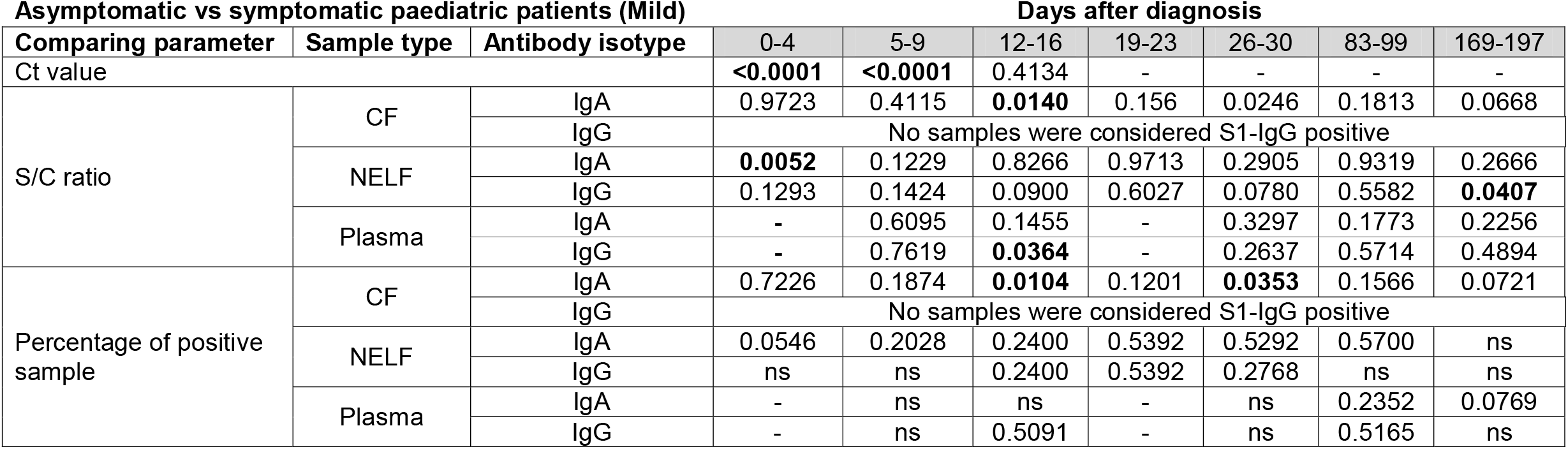
Comparisons of the SARS-CoV-2 S1-specific antibody levels between asymptomatic vs symptomatic paediatric patients with mild disease. The antibody levels in S/C ratios at the same time point were compared by Mann-Whitney test, while the percentages of positive sample were compared by Fisher’s Exact test. *P* values smaller than 0.05 are bolded, *p* values >0.9999 are represented by ns (not significant) while dashes means no data for comparisons.

| Asymptomatic vs symptomatic paediatric patients (Mild) |  |  | Days after diagnosis |  |  |  |  |  |  |
| --- | --- | --- | --- | --- | --- | --- | --- | --- | --- |
| Comparing parameter | Sample type | Antibody isotype | 0-4 | 5-9 | 12-16 | 19-23 | 26-30 | 83-99 | 169-197 |
| Ct value |  |  | **<0.0001** | **<0.0001** | 0.4134 | - | - | - | - |
| S/C ratio | CF | IgA | 0.9723 | 0.4115 | **0.0140** | 0.156 | 0.0246 | 0.1813 | 0.0668 |
|  |  | IgG | No samples were considered S1-IgG positive |  |  |  |  |  |  |
|  | NELF | IgA | **0.0052** | 0.1229 | 0.8266 | 0.9713 | 0.2905 | 0.9319 | 0.2666 |
|  |  | IgG | 0.1293 | 0.1424 | 0.0900 | 0.6027 | 0.0780 | 0.5582 | **0.0407** |
|  | Plasma | IgA | - | 0.6095 | 0.1455 | - | 0.3297 | 0.1773 | 0.2256 |
|  |  | IgG | - | 0.7619 | **0.0364** | - | 0.2637 | 0.5714 | 0.4894 |
| Percentage of positive sample | CF | IgA | 0.7226 | 0.1874 | **0.0104** | 0.1201 | **0.0353** | 0.1566 | 0.0721 |
|  |  | IgG | No samples were considered S1-IgG positive |  |  |  |  |  |  |
|  | NELF | IgA | 0.0546 | 0.2028 | 0.2400 | 0.5392 | 0.5292 | 0.5700 | ns |
|  |  | IgG | ns | ns | 0.2400 | 0.5392 | 0.2768 | ns | ns |
|  | Plasma | IgA | - | ns | ns | - | ns | 0.2352 | 0.0769 |
|  |  | IgG | - | ns | 0.5091 | - | ns | 0.5165 | ns |

**Figure 3.**
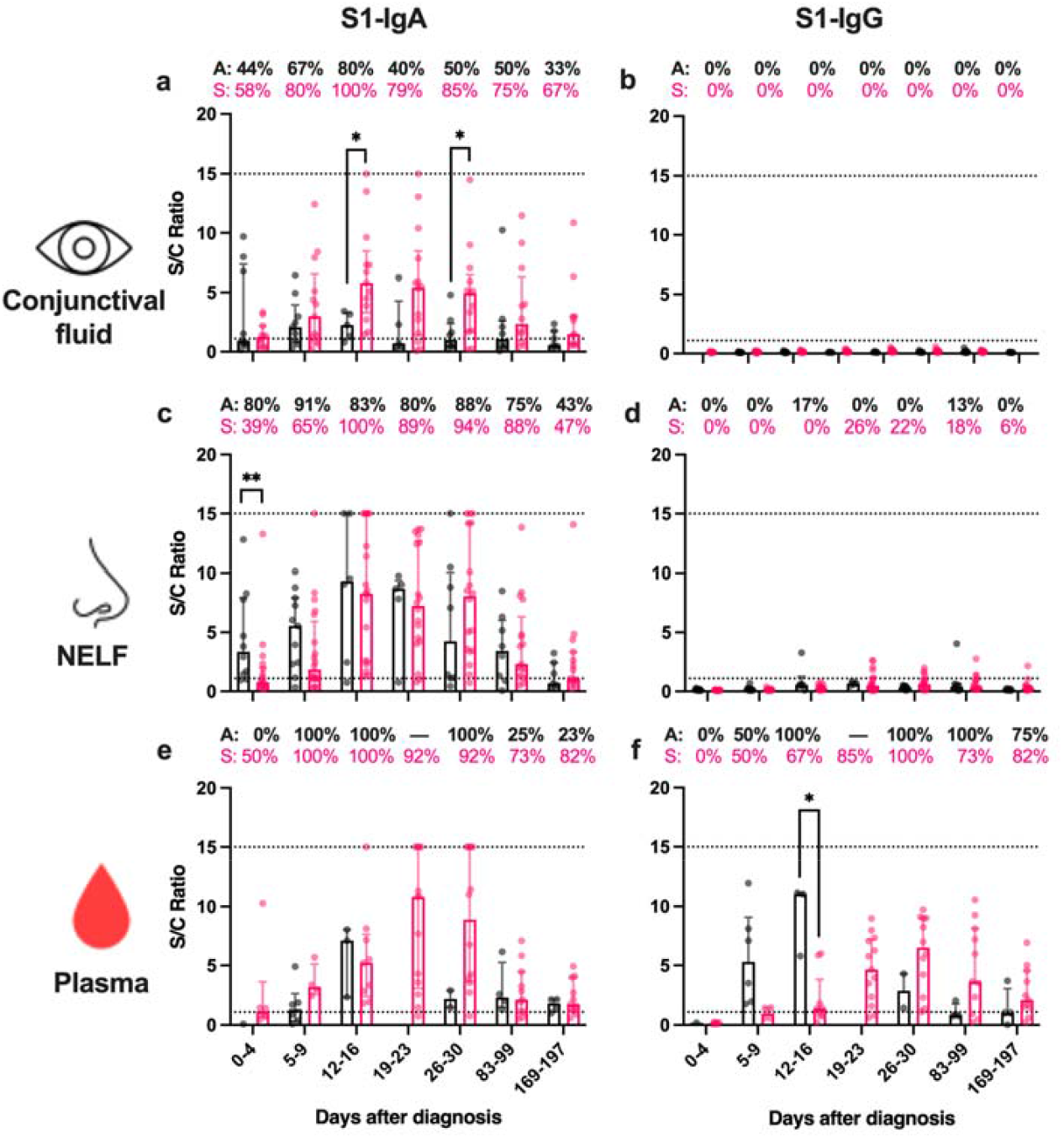
**SARS-CoV-2 S1-specific antibody levels in the (a-b) conjunctival fluid, (c-d) nasal epithelial lining fluid (NELF) and (e-f) plasma of in asymptomatic and symptomatic paediatric patients**. Grey and pink symbols indicate data of asymptomatic and symptomatic patients, respectively. Data points above the dotted line (Sample/Calibrator (S/C) ratio ≥ 1.1) are considered as positive, while the dotted lines at y=15 indicate the upper detection limit of the assay. The percentages denote the IgA and IgG positivity at each time point and the dash represents no data available. Median and interquartile range are plotted with dots represent individual value. The levels of SARS-CoV-2 S1-specific Ig were compared between asymptomatic and symptomatic patients by Mann-Whitney test. The asterisks indicate the statistical differences found, *: *p* <0.05 and **: *p* < 0.01.

### Asymptomatic paediatric patients had early induction of S1-specific IgA in their nasal mucosa and a higher level of S1-specific IgG in their plasma

In NELF of asymptomatic paediatric patients, there were significantly higher levels of S1-specific IgA (*p* = 0.0052, **Figure 3d**) and a trend of a higher percentage of positive S1-specific IgA on 0-4 days post-diagnosis (*p* = 0.0546). Moreover, a significantly higher level of S1-specific IgG was detected in the plasma of the asymptomatic than symptomatic paediatric subjects on 12-16 days post-diagnosis (*p* = 0.0364, **Figure 3f**).

### A higher percentage of NELF in patients with mild disease are S1-specific IgA positive by two weeks post-diagnosis

In adults, patients with mild disease had a higher percentage of NELF S1-specific IgA than those of the severe & critically ill group by 12-16 days post-diagnosis (*p* = 0.0106, **Table 2**) though there were no differences in the antibody levels (**Figure 4a**). In contrast, by 19-23 days post-diagnosis, severe & critically ill patients had significantly higher NELF S1-specific IgG levels than the mild patients (*p* = 0.0176, **Figure 4b**).

**Table 2.** Comparisons of the SARS-CoV-2 S1-specific antibody levels between adult patients of different disease severity. The antibody levels in S/C ratios at the same time point were compared by Mann-Whitney test, while the percentages of positive sample were compared by Fisher’s Exact test. *P* values smaller than 0.05 are bolded, *p* values >0.9999 are represented by ns (not significant) while dashes means no data for comparisons.

| Adult patients of different severity |  |  | Severity comparison | Days after diagnosis |  |  |  |  |  |  |
| --- | --- | --- | --- | --- | --- | --- | --- | --- | --- | --- |
| Comparing parameters | Sample type | Antibody isotype |  | 0-4 | 5-9 | 12-16 | 19-23 | 26-30 | 83-99 | 169-197 |
| Ct value |  |  | 1 vs 2 | 0.9860 | 0.1731 | 0.2349 | - | - | - | - |
|  |  |  | 1 vs 3+4 | 0.3187 | **0.0015** | **<0.0001** | - | - | - | - |
|  |  |  | 2 vs 3+4 | 0.2296 | **0.0073** | 0.0986 | 0.3860 | - | - | - |
| S/C ratio | NELF | IgA | 1 vs 2 | 0.8210 | 0.7687 | 0.6778 | 0.0839 | ns | 0.4002 | ns |
|  |  |  | 1 vs 3+4 | 0.3610 | 0.2419 | ns | 0.4380 | 0.9830 | ns | ns |
|  |  |  | 2 vs 3+4 | ns | ns | ns | ns | ns | ns | 0.9295 |
|  |  | IgG | 1 vs 2 | ns | ns | ns | 0.5200 | ns | ns | ns |
|  |  |  | 1 vs 3+4 | 0.0751 | **0.0207** | ns | **0.0176** | ns | ns | 0.1750 |
|  |  |  | 2 vs 3+4 | 0.3243 | **0.0379** | ns | 0.4394 | ns | ns | 0.4965 |
|  | Plasma | IgA | 1 vs 2 | ns | 0.2619 | ns | - | - | - | - |
|  |  |  | 1 vs 3+4 | ns | **0.0449** | 0.4238 | - | - | - | - |
|  |  |  | 2 vs 3+4 | ns | 0.8966 | ns | - | - | - | - |
|  |  | IgG | 1 vs 2 | 0.5996 | 0.8344 | 0.7138 | - | - | - | - |
|  |  |  | 1 vs 3+4 | 0.8856 | ns | ns | - | - | - | - |
|  |  |  | 2 vs 3+4 | ns | ns | 0.1636 | - | - | - | - |
| Percentage of positive sample | NELF | IgA | 1 vs 2 | 0.3845 | 0.7281 | 0.0877 | ns | ns | 0.387 | ns |
|  |  |  | 1 vs 3+4 | ns | 0.6758 | **0.0106** | ns | ns | 0.6499 | 0.6193 |
|  |  |  | 2 vs 3+4 | ns | ns | ns | ns | ns | ns | ns |
|  |  | IgG | 1 vs 2 | ns | ns | 0.4762 | 0.2105 | ns | ns | ns |
|  |  |  | 1 vs 3+4 | ns | ns | ns | 0.0686 | 0.5765 | 0.4706 | ns |
|  |  |  | 2 vs 3+4 | ns | ns | ns | ns | ns | ns | ns |
|  | Plasma | IgA | 1 vs 2 | 0.4375 | 0.1414 | ns | ns | - | - | - |
|  |  |  | 1 vs 3+4 | ns | 0.0699 | ns | - | - | - | - |
|  |  |  | 2 vs 3+4 | ns | 0.5147 | ns | - | - | - | - |
|  |  | IgG | 1 vs 2 | ns | ns | 0.4857 | ns | - | - | - |
|  |  |  | 1 vs 3+4 | ns | ns | ns | - | - | - | - |
|  |  |  | 2 vs 3+4 | ns | 0.5147 | 0.5238 | - | - | - | - |

**Figure 4.**
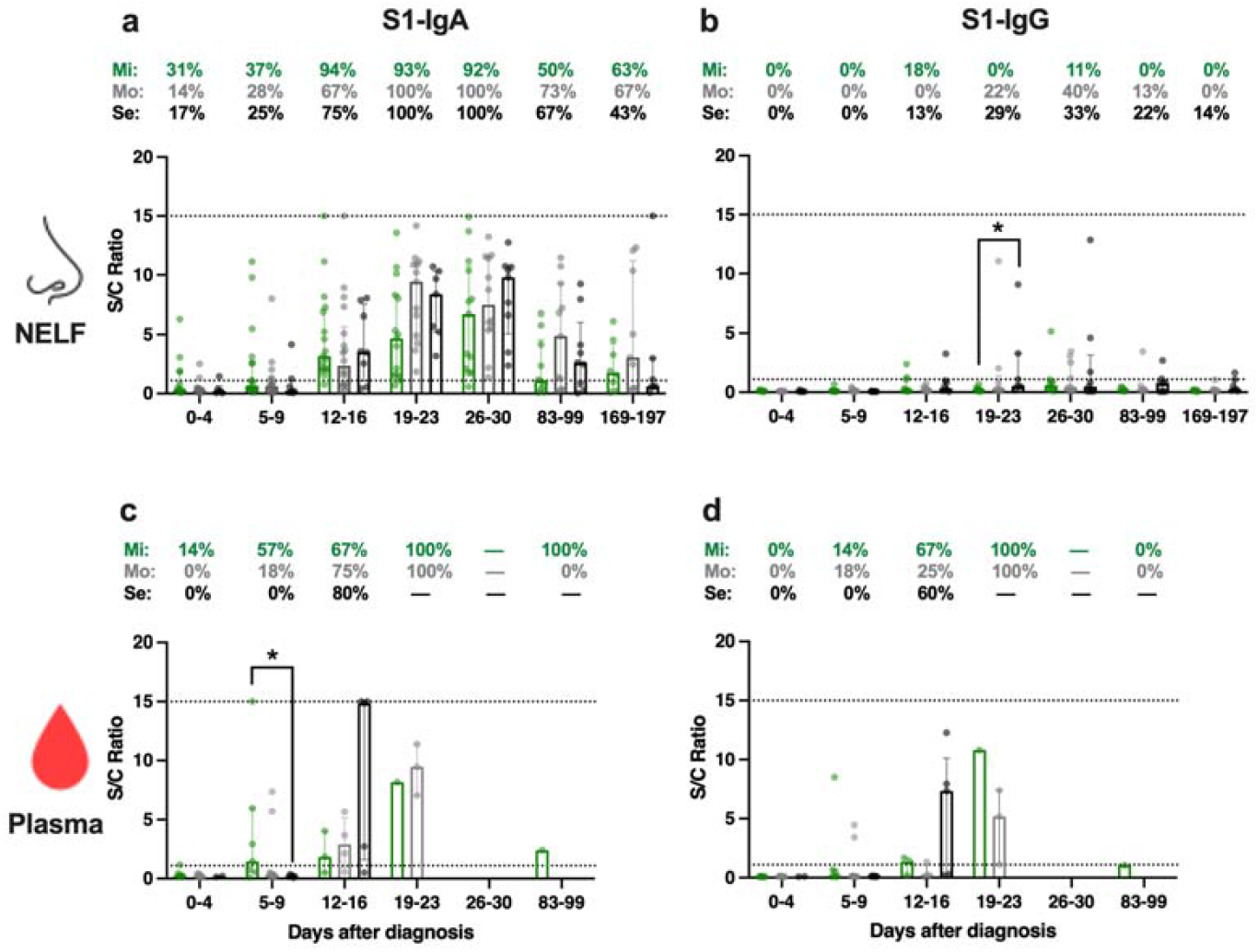
**SARS-CoV-2 S1-specific antibody levels in the (a-b) nasal epithelial lining fluid (NELF) and (d-e) plasma in adult COVID-19 patients of different disease severity from acute infection to convalescent phase**. Green, grey and black symbols indicate data of mild, moderate and severe & critically ill patients, respectively. Data points above the dotted line (Sample/Calibrator (S/C) ratio ≥ 1.1) are considered as positive, while the dotted lines at y=15 indicate the upper detection limit of the assay. The percentages denote the IgA and IgG positivity at each time point and the dash represents no data available. Median and interquartile range are plotted with dots represent individual value. The levels of SARS-CoV-2 S1-specific Ig were compared among disease severity groups by Kruskal-Wallis test followed by Dunn’s multiple comparisons test at each time point. The asterisks indicate the statistical differences found, *: *p* <0.05.

### Early induction of S1-specific IgA in the plasma of adult patients with mild disease

With the limited number of plasma samples collected, we focused our analysis on the first two weeks post-diagnosis. There were significantly higher levels of plasma S1-specific IgA (*p* = 0.0449, **Figure 4c**) and a trend of higher S1-specific IgA positivity (*p* = 0.0699) in patients with mild disease than those with severe or critical illness on 5-9 days post-diagnosis. In contrast, there were no statistically significant differences in the S1-specific IgG levels and the percentage of positivity in plasma among the severity groups at any time points (**Figure 4d, Table 2**).

### Paediatric patients had a higher level of S1-specific antibodies in their NELF and plasma during the early phase of SARS-CoV-2 infection than adult patients with mild disease

As 33 out of 34 paediatric patients had mild disease, we compared the mucosal and serological antibody responses between paediatric and adult patients (n=18) who had mild disease. Paediatric patients had significantly higher levels of NELF S1-specific IgA on 0-4, 5-9 and 12-16 days post-diagnosis (*p* = 0.0357, 0.0299 and 0.042, respectively). Moreover, a higher percentage of S1-specific IgA positive NELF was detected in paediatric patients than in adult patients on days 5-9 post-diagnosis (*p* = 0.04, Fisher’s exact test).

Paediatric patients also had a higher plasma level of S1-specific IgA (*p* = 0.0079) and IgG (*p* = 0.0333) on 0-4 days post-diagnosis and a trend of higher percentages of S1-specific IgA positive plasma samples on 0-4 (*p* = 0.0907), 5-9 (*p* = 0.0174) and 12-16 days post-diagnosis (*p* = 0.0384) than in adult patients with mild disease (**Table 3**). No differences in the percentage of S1-specific IgG positive plasma were found between the two age groups.

**Table 3.** Comparisons of the SARS-CoV-2 S1-specific antibody levels between paediatric and adult patients with mild disease. The antibody levels in S/C ratios at the same time point were compared by Mann-Whitney test, while the percentages of positive sample were compared by Fisher’s Exact test. *P* values smaller than 0.05 are bolded, *p* values >0.9999 are represented by ns (not significant) while dashes means no data for comparisons.

| Paediatric vs adult patients with mild disease |  |  | Days after diagnosis |  |  |  |  |  |  |
| --- | --- | --- | --- | --- | --- | --- | --- | --- | --- |
| Comparing parameter | Sample type | Antibody isotype | 0-4 | 5-9 | 12-16 | 19-23 | 26-30 | 83-99 | 169-197 |
| Ct value |  |  | **<0.0001** | 0.0601 | 0.7159 | - | - | - | - |
| S/C ratio | NELF | IgA | **0.0357** | **0.0299** | **0.042** | 0.0874 | 0.6640 | 0.1514 | 0.7303 |
|  |  | IgG | 0.8145 | 0.0981 | 0.2560 | **0.0392** | 0.7469 | **0.0069** | **0.0314** |
|  | Plasma | IgA | **0.0079** | 0.1791 | 0.0703 | - | - | - | - |
|  |  | IgG | **0.0333** | 0.1309 | 0.5363 | - | - | - | - |
| Percentage of positive sample | NELF | IgA | 0.2000 | **0.0400** | ns | ns | ns | 0.1068 | 0.6817 |
|  |  | IgG | ns | ns | 0.2269 | 0.2911 | ns | 0.5500 | ns |
|  | Plasma | IgA | 0.0907 | **0.0174** | **0.0384** | - | - | - | - |
|  |  | IgG | ns | 0.3156 | ns | - | - | - | - |

### Asymptomatic paediatric patients had a lower viral load during admission and adult patients with mild disease had a sharp reduction in viral load in the first week after disease diagnosis

Asymptomatic paediatric subjects had a significantly lower viral load than the symptomatic paediatric patients on 0-4 (*p* < 0.0001) and 5-9 days post-diagnosis (*p* < 0.0001) (**Figure 5a and Table 1**) while in adults, similar viral loads were detected among the three severity groups during admission (**Figure 5b and Table 2**).

**Figure 5.**
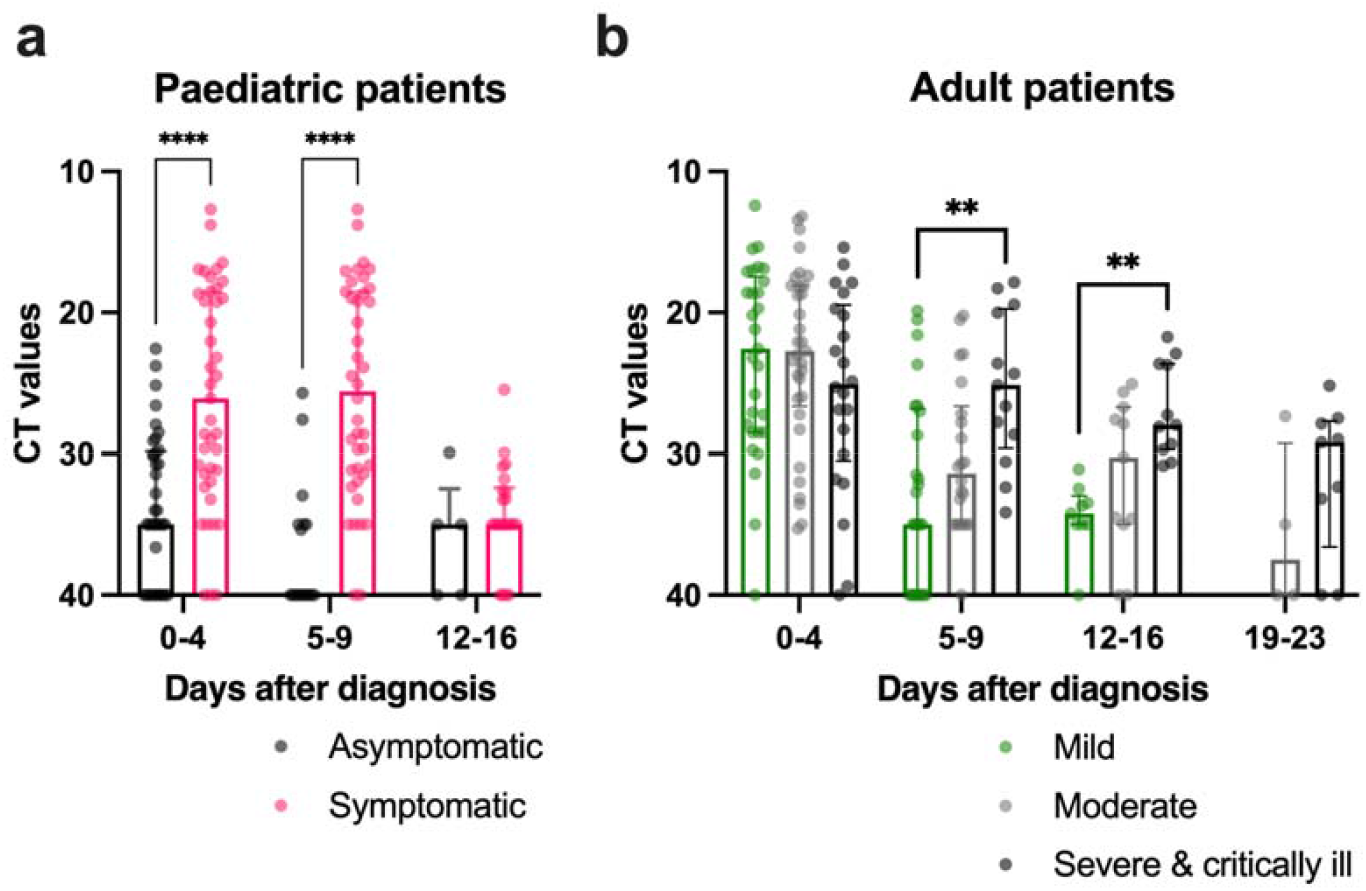
Comparison of SARS-CoV-2 viral load in (a) paediatric and (b) adult COVID-19 paediatric patients of different disease severity during hospitalization. The cycle threshold (CT) values of the SARS-CoV-2 viral gene in (a) asymptomatic and symptomatic paediatric patients were compared by Mann-Whitney test and among (b) mild, moderate and severe & critically ill adult patients by Kruskal-Wallis test followed by Dunn’s multiple comparisons test at each time point. Median and interquartile range are plotted with dots represent individual value. The asterisks indicate the statistical differences found, **: *p* < 0.01 and ****: p<0.0001.

Viral loads in asymptomatic paediatric patients decreased from 0-4 (median CT=33.59) to 5-9 (median CT= 40.00) days post-diagnosis (*p* = 0.0564, **Table 4a**). In comparison, viral loads of the symptomatic paediatric patients decreased later at 12-16 days post-diagnosis (median CT= 34.32), **Table 4b**. In adults, viral loads of patients in the mild and moderate groups declined over the three-week period while the viral loads in the severe and critically ill patients remained similar from 0-4 to 19-23 days post-diagnosis (**Table 4e**). As patients with mild disease were discharged earlier, no CT values were available in the subsequent timepoints for comparison.

**Table 4.**
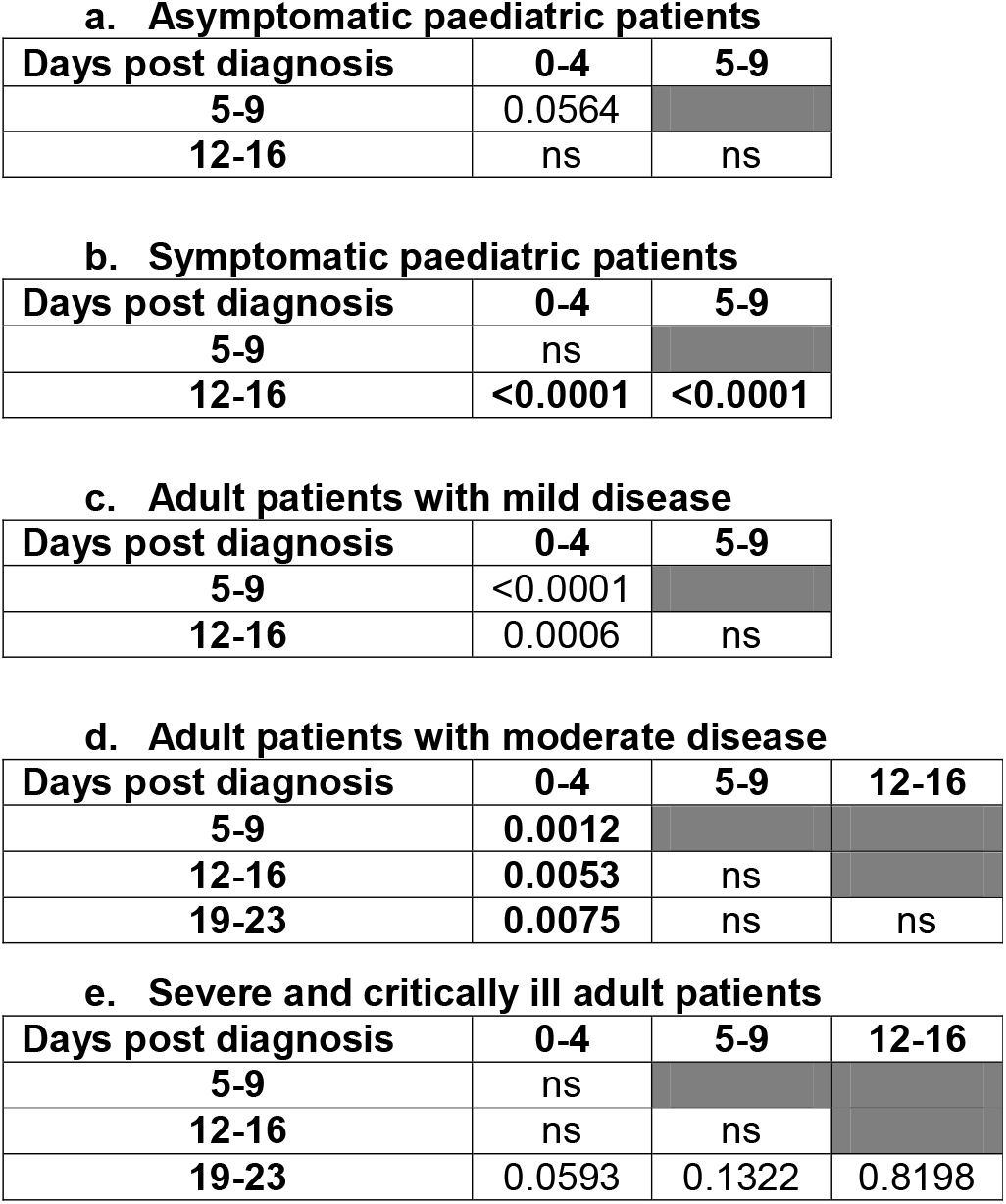
Comparisons of the Ct values of SARS-CoV-2 along time points. The ct values of the SARS-CoV-2 viral gene were compared by Kruskal-Wallis test across the time points followed by Dunn’s multiple comparison test. *P* values smaller than 0.05 are bolded, *p* values >0.9999 are represented by ns (not significant).

**a. Asymptomatic paediatric patients**
| Days post diagnosis | 0-4 | 5-9 |
| --- | --- | --- |
| 5-9 | 0.0564 |  |
| 12-16 | ns | ns |

**b. Symptomatic paediatric patients**
| Days post diagnosis | 0-4 | 5-9 |
| --- | --- | --- |
| 5-9 | ns |  |
| 12-16 | **<0.0001** | **<0.0001** |

**c. Adult patients with mild disease**
| Days post diagnosis | 0-4 | 5-9 |
| --- | --- | --- |
| 5-9 | **<0.0001** |  |
| 12-16 | 0.0006 | ns |

**d. Adult patients with moderate disease**
| Days post diagnosis | 0-4 | 5-9 | 12-16 |
| --- | --- | --- | --- |
| 5-9 | **0.0012** |  |  |
| 12-16 | **0.0053** | ns |  |
| 19-23 | **0.0075** | ns | ns |

**Table 4. Comparisons of the Ct values of SARS-CoV-2 along time points.**
| Days post diagnosis | 0-4 | 5-9 | 12-16 |
| --- | --- | --- | --- |
| 5-9 | ns |  |  |
| 12-16 | ns | ns |  |
| 19-23 | 0.0593 | 0.1322 | 0.8198 |

### Paediatric patients had lower viral loads than adult patients within the first week of diagnosis

When the viral loads of paediatric and adult patients with mild disease were compared, it was noted that initial viral loads of the asymptomatic paediatric patients were lower than corresponding adults (*p* < 0.0001, **Table 3**) and the viral loads in the paediatric patients were lower than adult subjects on 5-9 days post-diagnosis (*p* < 0.05).

### S1-specific IgA levels correlated positively with the SARS-CoV-2 neutralizing effect of the mucosal and plasma samples

53% (19/36) of CF had neutralizing effect. All these nineteen CF samples were S1-specific IgA positive. However, S1-specific IgA positivity did not translate directly to neutralizing effect, 37% (15/34) of S1-specific IgA positive CF did not inhibit the binding of angiotensin-converting enzyme (ACE) with the SARS-CoV-2 receptor binding domain (RBD). Nevertheless, higher levels of S1-specific IgA were detected in the neutralizing (median S/C ratio = 7.926) than the non-neutralizing CF samples (median S/C ratio =2.635, *p* = 0.0012). A significant positive correlation was found between the S1-specific IgA levels in CF (*p* = 0.0003, **Figure 6a**) and neutralizing antibody (NAb) levels.

**Figure 6.**
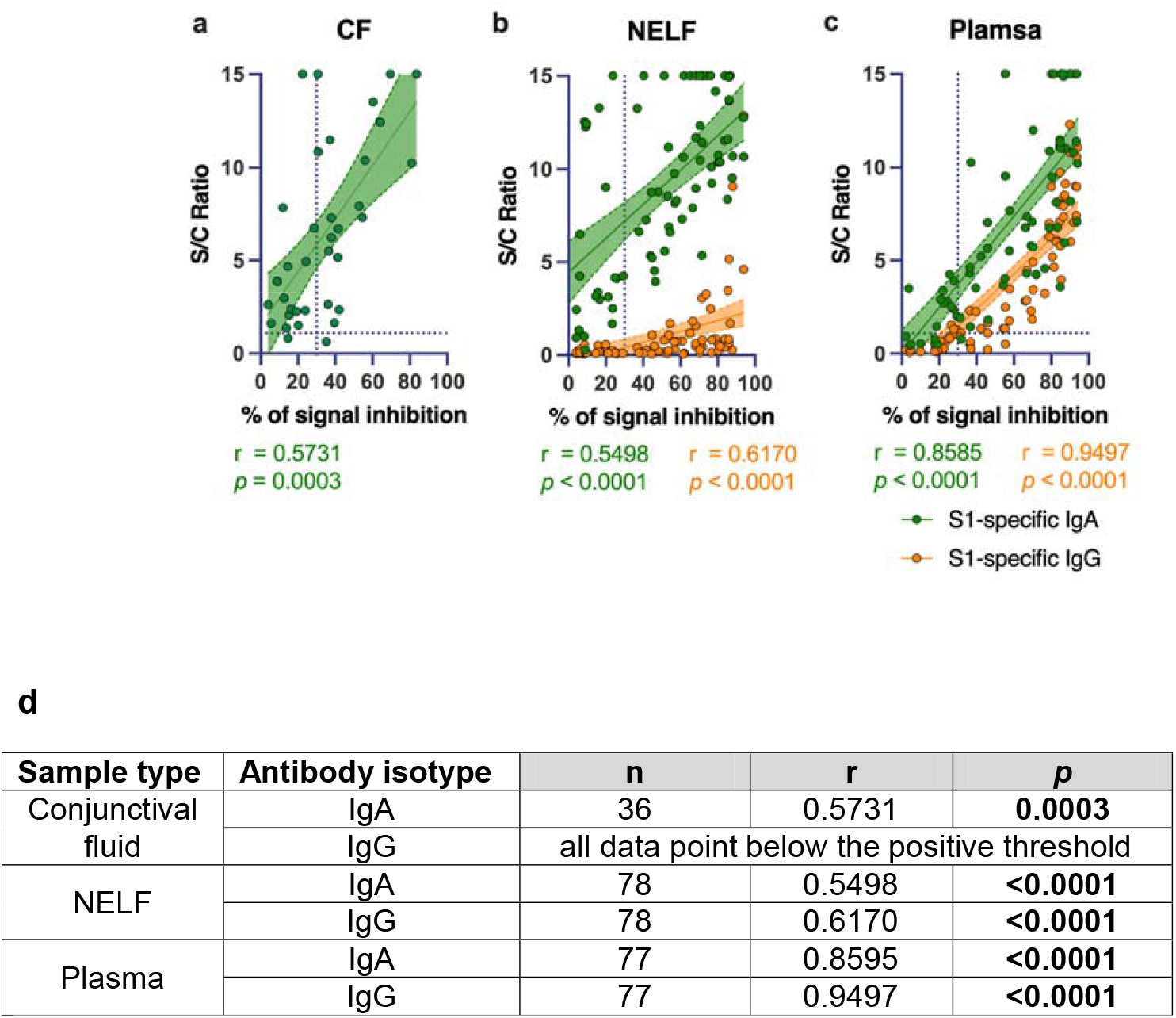
Correlation of SARS-CoV-2 S1-specific Igs to the percentage of signal inhibition in the surrogate ACE-2 based neutralization readout. The correlation coefficients of the **(a)** conjunctival fluid, **(b)** NELF and **(c)** plasma of COVID-19 patients are superimposed on the panel with trend lines estimated with the use of simple linear regression. Plots show the S/C ratio of the SARS-CoV-2 S1-specific IgA 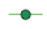 and IgG 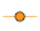 plotted against the percentage of inhibition of the SARS-CoV-2 Spike-ACE-2 binding signal, in which an inhibition ≥ 30% is regarded as the threshold of a positive sample, indicated by the vertical dotted line. Green and orange dotted lines represent significant linear regression fits with 95% confidence intervals (shaded region with the corresponding colours). **(d)** Table shows the number of sample included (n), the spearman r and the *p* value of the two-tailed test were shown. *P* values smaller than 0.05 are bolded.

68% (53/78) of NELF had neutralizing effect, all these fifty-three NELF samples were also S1-specific IgA positive. In comparison, only 25% (13/53) of the NAb positive samples were also S1-specific IgG positive. Higher levels of S1-specific IgA were detected in the neutralizing (median S/C ratio = 10.65) than the non-neutralizing NELF samples (median S/C ration = 4.140, *p* = 0.0002). Significant positive correlations were found between the S1-specific IgA (*p* < 0.0001, **Figure 6b)** and IgG levels (*p* < 0.0001, **Figure 6b)** with the NAb level in the NELF samples.

68% (53/77) of the plasma samples were SARS-CoV-2 neutralizing. 98% (52/53) and 85% (45/53) of the neutralizing plasma were S1-specific IgA and IgG positive, respectively. In contrast to NELF, it is worth noting that a high percentage of neutralizing plasma samples, 87% (46/53), were positive for both S1-specific IgA and IgG. Significant positive correlations were found between the S1-specific IgA (r = 0.8585; *p* < 0.0001), and IgG levels (r = 0.9497; *p* < 0.0001) with the NAb level in the plasma samples (**Figure 6c**).

### ‘Neutralizing’ S1-specific IgA in NELF detected in the first week of diagnosis correlated with a rapid decrease in viral load

Using the fixed-effect model, we showed that early induction of NELF S1-specific IgA was related to the rapid decrease in viral load. Paediatric patients who had any of their NELF S1-specific IgA level above the thresholds level of 4.386 in the first week after diagnosis had a more rapid decline in the viral load than those did not (*p* = 0.002, **Supplementary Figure 1b**).

## Discussions

SARS-CoV-2 can enter the body via inhalation or by self-inoculation directly to mucosal surfaces, especially when hand hygiene is inadequate. While it is known that mucosal IgA plays a vital role in the first line of defense against pathogens, including SARS-CoV-2,^28^ the kinetics of the ocular and nasal mucosal specific-IgA responses remain poorly defined. This study recruited paediatric and adult COVID-19 patients and profiled their SARS-CoV-2 S1-specific mucosal antibody levels longitudinally from hospital admission to six months post-diagnosis. The timing of the SARS-CoV-2 antibodies production, immunoglobulin isotypes, concentrations, neutralizing potency, antibody longevity, and relevance in different age groups were studied.

The evidence from serological studies suggests that the induction of SARS-CoV-2 specific antibodies is positively associated with the disease severity.^7, 8^ A study comparing the SARS-CoV-2 specific IgM, IgG and NAb in the sera of asymptomatic patients with sex-, age- and comorbidity-matched mild symptomatic patients during the acute and early convalescent phases identified that significantly lower IgG levels were detected in asymptomatic patients at both time points. There was also a quick decline of NAb levels in the early convalescent phase.^29^ Meanwhile, mucosal SARS-CoV-2–specific IgA response can be detected in subjects with minimal SARS-CoV-2 exposure, which could be accompanied by the absence of systemic virus-specific IgA production.^6^ The current longitudinal profiling of the antibodies reveals the IgA dominant mucosal response in COVID-19 patients.

In a meta-analysis, the prevalence of ophthalmic manifestations in patients with COVID-19 was 5.5%,^30^ though none of our paediatric patients had clinical symptoms or signs suggestive of conjunctivitis. Moreover, an animal study using rhesus macaques shows that conjunctival inoculation of SARS-CoV-2 virus would result in a less severe disease phenotype.^31^ Here, we studied the SARS-CoV-2 S1-specific antibody level in CF in paediatric subjects and elucidated the involvement of the conjunctival mucosa. The principal defense of the ocular mucosa is the secretory IgA (sIgA). The source of sIgA is from the lacrimal gland, where plasma cell and polymeric immunoglobulin receptor (pIgR) are present.^11^ IgG is not detectable in CF as the anterior chamber associated immune deviation of the eye tends to eliminate B cell which produces complement-fixing antibody, e.g. IgG.^32^ In addition, the CALT consists of central accumulations of IgA, IgD and IgM but not IgG-expressing CD20+ B cells.^33^ It is also described that not all healthy individuals have the CALT. The development of CALT starts in childhood and increases by the age of 10 years and subsequently declines with age.^34^ The surge of SARS-CoV-2 S1-specific IgA from 0-4 days to 12-16 days post-diagnosis in the symptomatic patients and the significantly higher S1-specific IgA levels in symptomatic than asymptomatic patients during the 2nd and 4th week post-diagnosis infer a more intense involvement of CALT in the symptomatic paediatric patients. Moreover, 67% of the CF from symptomatic patients remain S1-specific IgA positive by six months post-diagnosis, compared with 33% in the asymptomatic children.

The S1-specific IgA response is compartmentalized. When we assessed the antibody response in the nasal mucosa, an opposite pattern was observed. In the first four days of diagnosis, a more prominent S1-specific IgA was detected in the asymptomatic paediatric patients. While the NELF S1-specific IgA level correlated with the NELF neutralizing potency in the surrogate neutralization assay, the biological correlate of the NELF S1-specific IgA was further investigated. With the detection of ‘neutralizing’ NELF IgA in the first week after diagnosis, a more rapid viral load decline was observed. This pattern supports the hypothesis that the early involvement of SARS-CoV-2 specific-IgA limits the replication of SARS-CoV-2, thus the mild to asymptomatic disease presentation. It also infers that vaccines specialized in inducing nasal immune responses and memory cells could be of a remarkable advantage over those that only induce circulating antibody responses.

The mucosal S1-specific IgA response was localized, age-dependent, and correlated weakly with plasma IgA antibody levels. An early and robust NELF S1-specific IgA was induced in paediatric patients rather than adult patients with mild disease. Paediatric patients with mild disease had higher NELF S1-specific IgA levels than adult patients from 0 to 16 days post-diagnosis. Interestingly, infected children were found to have lower levels of spike and nucleocapsid antibodies in plasma than adults but a more expanded response to accessory proteins.^35^ In our study, all adult patients had low NELF S1-specific IgA levels on 0-4 days post-diagnosis, while the first statistically significant increase of S1-specific IgA in adult patients was detected only by 12-16 days post-diagnosis. Moreover, there were no differential S1-specific IgA levels in NELF among patients of different disease severity at all time points. Nevertheless, more adult patients with mild disease developed NELF S1-specific IgA on 12-16 days post-diagnosis than those with severe or critical illness. Interestingly, there seems to be a higher percentage of adult patients with mild disease having detectable plasma S1-specific IgA rather than NELF on 5-to-9 days after diagnosis. However, the limited number of plasma samples restraint us to perform further statistical investigation.

The early production of a sufficient level of NELF S1-specific IgA infers protective effects, such as a shorter viral shedding period and a milder disease presentation, which is also true in the case of human 229E infection.^36^ In contrast, the CF and blood S1-specific IgA levels detected on or beyond 12 days post-diagnosis were associated with disease severity, together with the extended longevity of the mucosal and plasma antibodies as reported in other studies.^37, 38, 39^

From the diagnostic point of view, as the mucosal antibody readout may appear before symptoms occur and it can also pick up virus exposed individual who is negative in a serological test,^40^ the mucosal antibody would be a feasible screening parameter for detecting asymptomatic individuals, especially during the early stage of the disease. Indeed, NELF S1-specific IgA provides a sensitive readout as evidence of SARS-CoV-2 infection in the first month of disease diagnosis. All subjects were detected with NELF S1-specific IgA in at least one time point. However, the duration from a prior SARS-CoV-2 infection inevitably affects the sensitivity of both mucosal and serological antibody tests,^38^ though mucosal S1-specific IgA appears to be less affected.^19^ The NELF S1-specific IgA was still detectable in at least 50% of the COVID-19 patients after three months (day 83-99) of diagnosis. After all, the non-invasive nature and the validity of paper strips used in our study would allow repeated tests without blood sampling.^26, 41^

One of the major limitations of this study was that we determined the SARS-CoV-2 S1-specific antibody only in its IgA and IgG isotypes. The diversity of the antibody responses to other SARS-CoV-2 viral antigens (e.g. nucleocapsid, open reading frame (ORF)8 and ORF3b) were not evaluated. These non-neutralizing antibodies are found to be detectable in COVID-19 patients plasma early in the disease course and serve crucial functions to counteract the viral inhibition on host antiviral effects.^35^ It would be of exceptional importance to characterize the antibody diversity in the mucosal fluids, so as to better determine its immunological involvement of differing disease outcomes. Moreover, the limited number of severe & critically ill patients limited the statistical power of the analysis.

Another limitation of our study was the lack of a cell-based neutralization test or plaque reduction assay, which required Biosafety level 3 facilities. In addition, the direct measurement of S1-specific antibody might not confer immunity, while the surrogate neutralization assay is expensive and not affordable if the number of samples to be tested is enormous. Here, we derived a threshold of S1-specific IgA levels which is predicted to have the ‘neutralizing’ potency. We applied this threshold to the rest of the NELF S1-specific IgA measurements and converted it to a biologically rational observation. Patients with ‘neutralizing’ NELF within the first 7 days of diagnosis had a more rapid decline in viral load. This translational threshold might be a workable alternative for laboratories that lack the necessary facilities and diagnostic utilities.

In conclusion, the longitudinal study on the mucosal antibody kinetics in COVID-19 paediatric and adult patients draws the picture for the independence of mucosal and systemic response. The higher mucosal IgA levels seem protective and present in asymptomatic subjects or patients with mild disease. The high mucosal IgA level is in great contrast to our understanding of the antibody profile in the circulation, in which high antibody levels are linked with severity. The differential intensity of the secretory IgA between paediatric and adult patients should arouse more attention, at least in the characterization of the mucosal antibody spectrum and their implication to the clinical presentation. The current finding provides an extra dimension for the diagnostic use of the mucosal antibody measurements, especially for the individuals at their early phase of infection and asymptomatic patients.

## Methods

### Subject recruitment and Severity Scoring

The presence of SARS-CoV-2 infection was confirmed by two RT-PCR tests targeting different regions of the RdRp gene. Paediatric and adult patients hospitalized with COVID-19 were recruited prospectively, if they were within four days of their first RT-PCR positive result. Longitudinal biospecimen collections were conducted at seven-time points during the in-patient period and after discharge. Patients were ready for discharge when they consecutively tested negative for SARS-CoV-2 by RT-PCR or had a viral CT value above 32 and tested positive for N-specific serum IgG by chemiluminescent microparticle immunoassay assay. Biospecimens of paediatric patients were collected by healthcare workers during the follow-up appointments while adult patients’ biospecimens were obtained by self-collection (**Figure 1a**). Conjunctival fluid samples (CF), nasal epithelial lining fluid samples (NELF) and plasma samples were collected during the in-patient and follow up appointments, while NELF was collected in discharged adult patients by self-collection method. Disease severity of these subjects was categorized into mild (score 1, the clinical symptoms were light, and there was no sign of pneumonia on imaging), moderate (score 2, with fever, respiratory tract and other symptoms, imaging suggests pneumonia), severe (score 3, coincide with any of the following: (1) respiratory distress, respiration rate (RR) ≥ 30 times / min; (2) the oxygen saturation ≤ 93% in the resting state; (3) PaO_2_ / FiO_2_ ≤ 300 mmHg (1mmHg = 0.133 kPa)) and critically ill (score 4, coincide with any of the following: (1) respiratory failure occurs and mechanical ventilation is required; (2) shock; (3) the patient develops other organ failure and needs ICU monitoring and treatment), as described in WHO COVID-19 Clinical management Living guidance.^27^ The study was approved by the Joint Chinese University of Hong Kong – New Territories East Cluster Clinical Research Ethics Committee (CREC: 2020.076 and 2020.4421).

### Conjunctival fluid (CF) and nasal epithelial lining fluid (NELF) collection

Sampling was conducted on both eyes with a technique similar to Schirmer’s test. The ocular strip (I-Dew Tear Strips) was inserted into the lower conjunctival sac and collected after the fluid reached the 25mm mark. The nasal strip made of Leukosorb was inserted into each nostril after 100ul of sterile saline was instilled as described ^17, 20^ followed by a one-minute nose pinch. All strips were collected and transferred in a dry sterile collection tube and eluted within 24h after collection.

### Elution of CF and NELF and plasma collection

To elute, ocular or nasal strips were soaked in 300ul phosphate-buffered saline (PBS) on ice. The solution and the strips were transferred to a Costar Spin-X (CLS9301) and centrifuged at 4°C to elute the CF or NELF. 3ml of blood was collected by venepuncture and transferred into EDTA blood tube. Plasma samples were separated by centrifugation at 4°C, 2000g for 20 mins. The specimens were aliquoted into small volume vials for downstream analysis of SARS-CoV-2 specific Ig panels and neutralization test and stored at −20°C until analysis.

### Measurement of SARS-CoV-2 Spike protein specific IgA and IgG

Semi-quantitative measurements of SARS-CoV-2 Spike protein (S1 domain) specific Ig ELISA Kits (Euroimmun, EI 2606-9601 A and EI 2606-9601 G) were used. 1:10 diluted-CF and NELF and 1:100 diluted plasma were assayed as per manufacturer’s instructions and analyzed on the Synergy HTX Multi-Mode Reader. Semi-quantitative readout as a ratio between the sample and the calibrator optical density (OD) was used. Data was expressed in Sample/Calibrator (S/C) ratio, a value ≥ 1.1 was considered as positive.

### Measurement of SARS-CoV-2 neutralization antibody (NAb)

A blocking enzyme-linked immunosorbent assay (GenScript, L00847) was employed. Briefly, undiluted CF, NELF, 1:9 diluted plasma samples and controls were processed as per manufacturer’s instructions. Samples which gave signal inhibition of ≥ 30% were considered as SARS-CoV-2 NAb positive.

### Statistical analysis

The demographic variables of subjects were compared between disease severity groups using Mann Whitney test, Kruskal-Wallis test and Fisher’s exact test, as appropriate. For the immunoglobulin profiles, differences between the different genders and age groups were evaluated using Mann Whitney test, while differences in disease severity groups and time points were tested using Kruskal-Wallis test followed by Dunn’s multiple comparison test. The correlation of S/C ratio of the specific immunoglobulins with the percentage of signal inhibition in the surrogate neutralization test was examined by Spearman’s correlation test. The estimation of the threshold level of NELF S1-specific to have neutralizing effect was determined by ROC curves using 78 NELF samples. Using the ROC curve, threshold was derived using the Youden Index calculation with the assumption that sensitivity and specificity hold equal diagnostic importance, *J* provides an optimal threshold.

*J* = Sensitivity + Specificity − 1, such that the threshold used provides maximum *J* The threshold provided the basis for predicting the neutralizing effect. When the S1-specific IgA S/C level was above the threshold, neutralizing effect of the NELF sample was predicted and vice versa. A fixed effects model was used to determine the differences in the rate of viral loads declined with time between samples from paediatrics patients who had neutralizing effect (estimated?) at 1st week of diagnosis or not. All statistical tests were performed using Graphpad version 9.1.2 for macOS SPSS version 25. Differences were considered statistically significant at *p* < 0.05.

## Data Availability

The Data will be available upon request.

## Acknowledgements

We would like to acknowledge Prof Aaron HP Ho, Prof Megan YP Ho and Miss Yuan-yuan Wei (Department of Biomedical Engineering, Faculty of Engineering, The Chinese University of Hong Kong) who tailor-cut the nasal strips for this study and Ms Fiona Cheng (Department of Paediatrics, The Chinese University of Hong Kong) for her assistance in preparing all the nasal strip vials. We would like to offer our special thanks to Ms Rity Wong and Ms Vickie Li (Department of Medicine and Therapeutics, The Chinese University of Hong Kong), Ms Angela Ho (Department of Ophthalmology and Visual Sciences, The Chinese University of Hong Kong), Ms Apple Yeung and Mr Lam Lap Yee (Department of Microbiology, The Chinese University of Hong Kong) for the assistant in the sample logistics. We thank all the subjects and their parents who agreed to participate in this study. We are grateful for Prof Ellis KL Hon’s (Department of Paediatrics, The Chinese University of Hong Kong) continuous support to the research team. This study is supported by the Health and Medical Research Fund (HMRF) commissioned grants COVID190112 (RWYC), HMRF commissioned grants COVID190103 (MHW), The Chinese University Direct Research Grant 2020.073 (RWYC), the Hong Kong Institute of Allergy Research Grant 2020 (RWYC) and the Innovation and Technology Fund PRP/039/21FX (RWYC).

## Author Contributions

Conceptualization: RWYC, KYYC, KKLC, AML, HSL

Methodology: RWYC, KCCC, GCYL, JGST, KYYC, KKLC, AML, HSL

Investigation: RWYC, JGST, KYYC, SL

Visualization: RWYC, KYYC, MHW

Funding acquisition: RWYC, MHW, PKSC

Project administration: RWYC, JGST, KYYC, AML, HSL,

Supervision: RWYC, KCCC, GCYL, KYYC, AML, HSL

Writing – original draft: RWYC, KCCC, JSKY

Writing – review & editing: RWYC, KCCC, GCYL, KYYC, MWLY, KKLC, RWYN, PKSC, AML, HSL

---

**Supplementary Figure 1.**
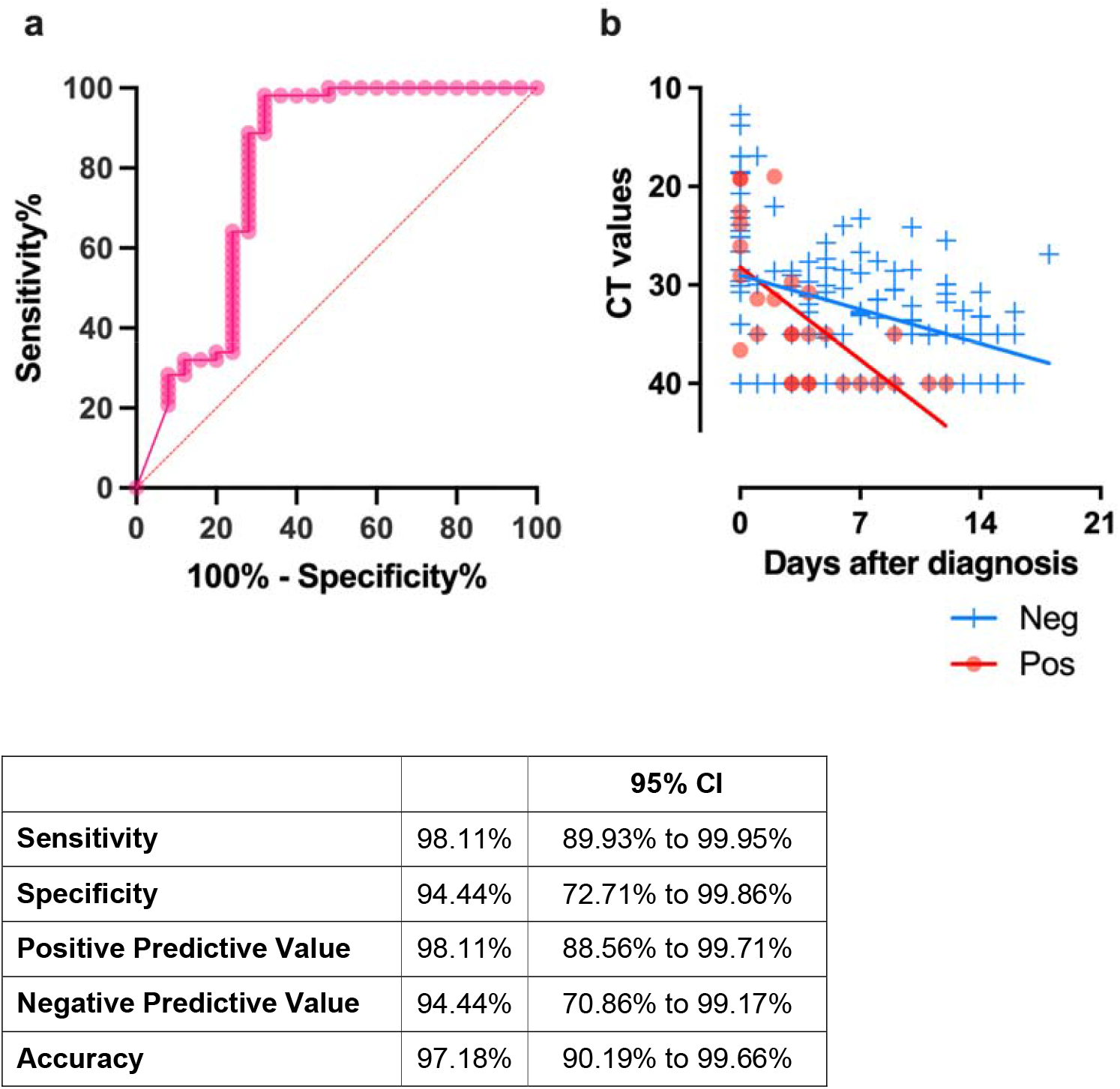
**(a)** Receiver operating characteristic (ROC) curves constructed using 78 NELF samples with both S1-specific IgA and NAb levels measured. The area under curve (AUC) was 0.80 with *p* < 0.001. Using the ROC curve, thresholds for NELF IgA was defined as > 4.386 by Youden Index calculation. Using this cutoff value, the sensitivity and specificity was 98.11% and 94.44%, respectively, with an accuracy of 97.18%. **(b)** All the available CT values of the paediatric patients who had any of their NELF S1-specific IgA level above 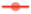 and below 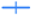 the thresholds level are plotted against time. A statistically significant difference was found in the decline rate of the viral load, *p* = 0.002.

